# Spatial heterogeneity of menstrual discriminatory practices against Nepalese women: a population-based study using the 2022 Demographic and Health Survey

**DOI:** 10.1101/2024.04.04.24305345

**Authors:** Geoffrey Barini, Sharon Amima, Damaris Mulwa, Polycarp Mogeni

**Affiliations:** Department of Pure and Applied Mathematics, Jomo Kenyatta University of Agriculture and Technology, Nairobi, Kenya; Department of Food Science, Nutrition and Technology, University of Nairobi, Nairobi, Kenya; Department of Statistics and Actuarial Sciences, Jomo Kenyatta University of Agriculture and Technology, Kenya; Kenya Medical Research Institute (KEMRI), Nairobi, Kenya

## Abstract

**Background:** Menstrual discrimination hinders progress toward Sustainable Development Goals including gender equality, health, education, and sanitation. Examining spatial heterogeneity of menstrual discriminatory practices presents an opportunity for targeted interventions. Our goal was to evaluate geographical disparities in menstrual restrictions among Nepalese women and assess their association with socio-economic and demographic factors.

**Methodology:** We utilized data from the 2022 Nepal Demographic and Health Survey, focusing on women aged 15 to 49 who had experienced a menstrual period in the last year. Our analysis explored the spatial patterns of menstrual restriction outcomes related to engagement in religious activities, household level restrictions, and the severe form of menstrual restrictions referred to *Chhaupadi*. The standard Gaussian kernel density estimation was used to create continuous prevalence maps and the spatial scan statistic was used to detect local clusters. The association between the different forms of menstrual restriction and the socio-economic, and demographic factors were examined using the Poisson regression model with robust standard errors.

**Findings:** Overall, the prevalence of women who reported any form of menstrual restriction was 84.8% and was subject to geographical variations ranging from 79.0% in Bagmati to 95.6% in Sudurpashchim. Religious restrictions were the most prevalent (79.8%) followed by household level restrictions (39.5%) and then *Chhaupadi* (6.2%). Geographical variations were detected in all forms of menstrual restriction outcomes and were more prominent for *Chhaupadi*. Individual level factors (age, sex, ethnicity/caste), household level factors (household wealth, age and sex of the household head) and regional factors (province) were significantly associated with at least one form of menstrual restriction outcome. Strikingly, women who reside in households led by female household heads were less likely to report experiencing household level restriction during menstruation (Adjusted prevalence ratio (aPR) =0.89 [95%CI: 0.84–0.94], p<0.001) whilst those residing in wealthy households were associated with lower likelihood of experiencing *Chhaupadi* (aPR =0.26 [95%CI: 0.17-0.39] among the richest).

**Interpretation:** We observed geographical micro-variations in menstrual discriminatory practices in Nepal presenting an opportunity for targeted behavioral interventions. Women empowerment is a promising strategy in the fight against menstrual discrimination within the household. Therefore, community-based targeted interventions should be designed and tested to determine their utility against *Chhaupadi* in these settings.

## Introduction

Although the last decade has witnessed significant investment in Water, Sanitation and Hygiene (WASH) programs, menstrual health and hygiene still pose a significant public health and social challenges in low- and middle-income countries (LMIC) (Hennegan et al., 2019; Kuhlmann et al., 2017; Plesons et al., 2021, 2023; UNICEF, 2022; Wood et al., 2022). Millions of women still encounter difficulties in managing their menstrual periods while preserving their dignity. This is predominantly attributed to internalized stigma and shame rooted in myths and misconceptions surrounding menstruation (ActionAid, 2024; Health, 2018; Health–Americas, 2022; Lonkhuijzen et al., 2023; UN, 2019; UNICEF, 2022). These myths and misconceptions, which regard menstrual blood as a contaminant are deeply rooted in the cultural and religious beliefs (Crawford et al., 2014; Health, 2018, 2018; Lonkhuijzen et al., 2023; Olson et al., 2022; Rothchild & Piya, 2020).

In the South Asian context, Nepal is one of the most affected countries, with over 90% of menstruating girls and women experiencing some form of discriminatory practices during their menstrual periods (UNICEF, 2020). These practices encompass restrictions on accessing places of worship, participation in household chores, physical contact with male household members, and constraints on utilizing water sources (ActionAid, 2024; Mukherjee et al., 2020; Rothchild & Piya, 2020; Thakuri et al., 2021). The western part of Nepal is known for the infamous *Chhaupadi*—the severest form of menstrual exclusion—that designates women as impure and untouchable during menstruation or postpartum period banishing them to a separate house, cowshed or menstrual hut (ActionAid, 2024; Amatya et al., 2018; Amery et al., 2023; Thakuri et al., 2021). Despite legislative measures against menstrual discriminatory practices, recent studies reveal that this practice is still pervasive in the western part of the country (Adhikari, 2020; Amatya et al., 2018; Baumann, Merante, et al., 2021; UNFPA, 2019; Vaughn, 2019), subjecting women to physical and mental health challenges, sexual abuse, snake bites and even death due to suffocation (Adhikari, 2020; Awasthi et al., 2022; Cardoso et al., 2019, 2019; Joshi & Acharya, 2022; Preiss, 2016; Thakuri et al., 2021).

Only recently has the global health and social research community endeavored to confront menstrual hygiene management as a substantial development concern and a barrier to achieving gender equality (Bobel, 2019; Health, 2018; Hennegan, 2019; Thomson et al., 2019; UN, 2019). The societal impact of menstrual hygiene mismanagement extends beyond adolescence, affecting adult women in their occupational engagements (Borg et al., 2023; Health–Americas, 2022; Hennegan et al., 2020; Krenz & Strulik, 2021; Sumpter & Torondel, 2013). Recent studies have described the association between menstrual hygiene management and poor school performance or attendance among girls, coinciding with the scale-up of WASH intervention programs in schools (Alam et al., 2017; Habtegiorgis et al., 2021; Sommer et al., 2015, 2016; Sumpter & Torondel, 2013; Tegegne & Sisay, 2014). Further progress is likely if more research is dedicated towards fighting the negative cultural believes and taboos surrounding menstrual hygiene management. Therefore, the development of risk stratification maps of menstrual discriminatory practices means that the most vulnerable populations can be identified and prioritized with scarce resources.

Using the 2022 Nepal Demographic and Health Survey (NDHS) dataset, we explored the spatial patterns of *a priori* defined menstrual restriction outcomes related to engagement in religious activities, performance of household activities/interactions within the household and *Chhaupadi*. The second objective was to assess the social-economic and demographic factors associated with the geographical variations in risk of menstrual discrimination. This is one of the few studies to describe the spatial heterogeneity of menstrual discriminatory practices in Nepal and the first to do so among women aged 15 – 49 years using a nationally representative population-based survey.

## Materials and Methods

### Ethics

The 2022 NDHS protocol was developed and reviewed by the Nepal Health Research Council and the ICF Institutional Review Board. Written consent from the household head was a prerequisite for conducting the interviews (Corsi et al., 2012). Here we conducted a secondary data analysis using de-identified data from the 2022 NDHS, which is publicly available and does not require ethical approvals. Permission to use the data was obtained through the DHS division at ICF International, following the completion of an online request form available on the DHS website (Corsi et al., 2012). Therefore, no ethical reviews or approvals were necessary for the preparation of the manuscript.

### Study site

Nepal is a landlocked country situated in southern Asia between latitudes 26° and 30° N and longitudes 80° and 89° E. Nestled along the southern slopes of the Himalayas, Nepal shares borders with India to the east, west, and south, and China to the north. Covering an area of 147,516 square kilometers, the country has a population of approximately 29.2 million people, with females comprising 51.1%. The country is divided into seven provinces: Koshi, Madhesh, Bagmati, Gandaki, Lumbini, Karnali, and Sudurpashchim. These provinces are subdivided into 77 districts, housing a total of 753 local-level municipalities, with 293 classified as urban and 460 as rural. Nepal’s topography encompasses three ecological zones: Terai (plains), hills, and mountains. Approximately 53.7% of the population resides in the Terai region, with 40.2% in the hilly areas and only 6.1% in the mountainous regions. Approximately, 65% of the population lives in urban centers. The country boasts a diverse culture, comprising over 120 castes/ethnicities, with Hinduism as the predominant religion (National Statistics Office, 2021).

### Study design and data source

The 2022 NDHS dataset is a nationally representative population-based survey that is freely available upon request (Corsi et al., 2012). Details of the study design, consenting, data collection, supervision and training procedures have been described elsewhere (ICF, 2023). Briefly, the 2022 NDHS survey adopted a two-stage stratified sampling methodology that included the segmentation of each of the seven provinces into urban and rural categories, yielding a total of 14 sampling strata. Within each stratum, the sampling frame was organized based on administrative units, and probability-proportional-to-size selection was applied in the first stage. In the initial sampling phase, 476 primary sampling units (PSUs) were chosen, with the probability proportional to PSU size and independent selection was done within each stratum. Out of these PSUs, 248 were from urban areas, and 228 were from rural areas. Prior to the main survey, a household listing exercise was conducted in all selected PSUs, establishing the sampling frame for the second-stage selection of sample households. A total of 14,280 households were selected, with thirty households chosen from each cluster. Household-level Global Positioning System (GPS) data were collected as part of the household listing process.

### Study participants

Our analyses are restricted to women and girls aged 15–49 years. Social and demographic characteristics of the study participants are summarized in the (**Supp. Table 1**).

**Table 1:**
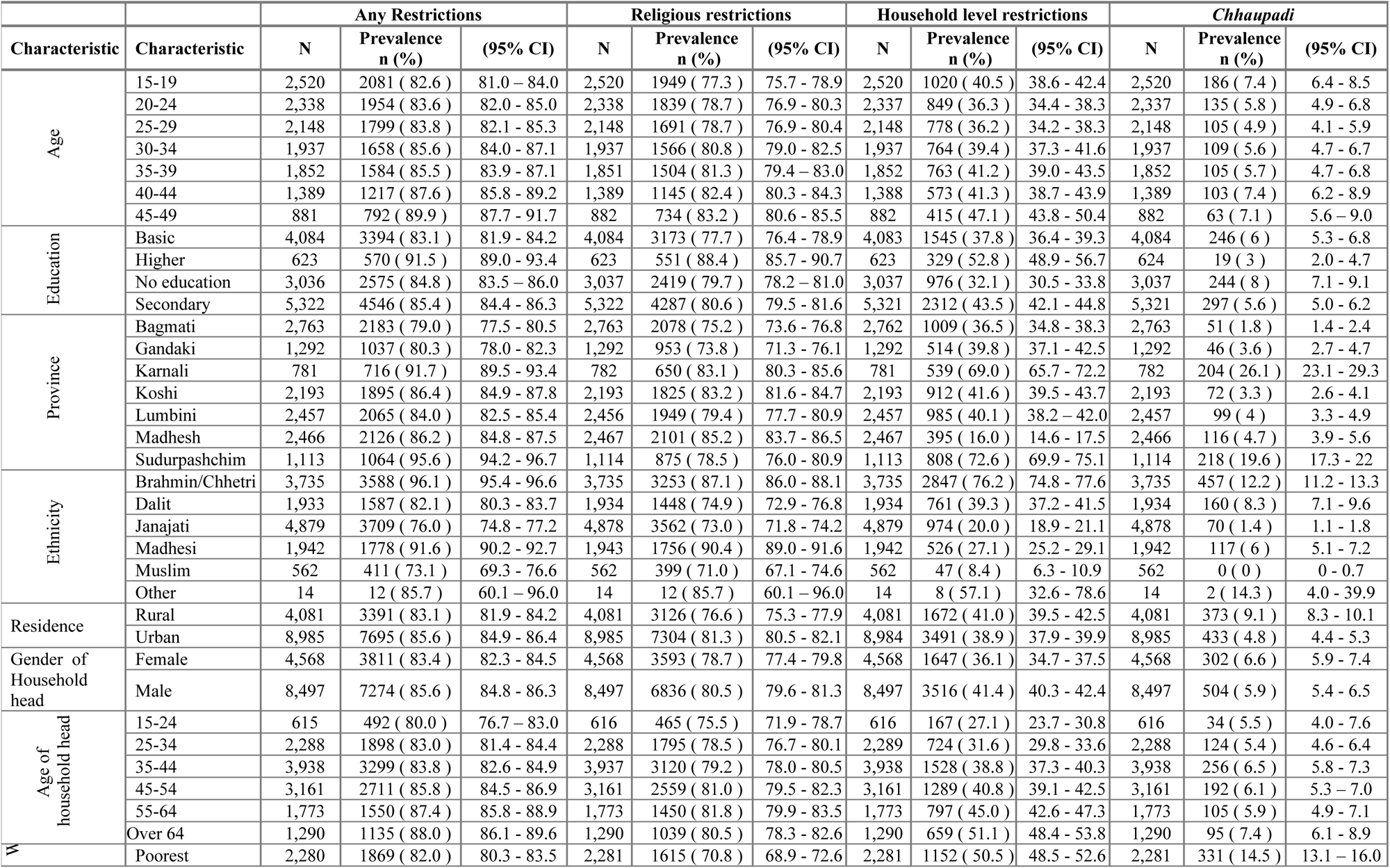
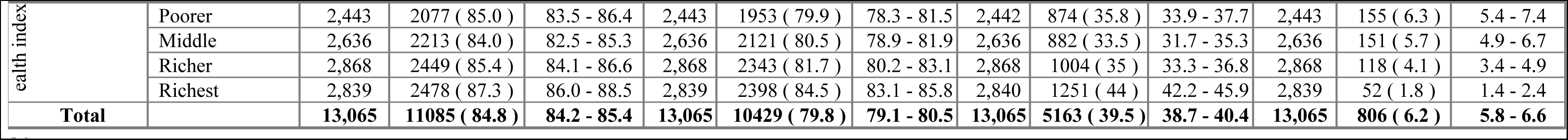
Prevalence of menstrual restrictions by social-demographic and economic factors.

|  |  | Any Restrictions |  |  | Religious restrictions |  |  | Household level restrictions |  |  | *Chhaupadi* |  |  |
| --- | --- | --- | --- | --- | --- | --- | --- | --- | --- | --- | --- | --- | --- |
| Characteristic | Characteristic | N | Prevalence<br>n (%) | (95% CI) | N | Prevalence<br>n (%) | (95% CI) | N | Prevalence<br>n (%) | (95% CI) | N | Prevalence<br>n (%) | (95% CI) |
| Age | 15-19 | 2,520 | 2081 ( 82.6 ) | 81.0 – 84.0 | 2,520 | 1949 ( 77.3 ) | 75.7 - 78.9 | 2,520 | 1020 ( 40.5 ) | 38.6 - 42.4 | 2,520 | 186 ( 7.4 ) | 6.4 - 8.5 |
|  | 20-24 | 2,338 | 1954 ( 83.6 ) | 82.0 - 85.0 | 2,338 | 1839 ( 78.7 ) | 76.9 - 80.3 | 2,337 | 849 ( 36.3 ) | 34.4 - 38.3 | 2,337 | 135 ( 5.8 ) | 4.9 - 6.8 |
|  | 25-29 | 2,148 | 1799 ( 83.8 ) | 82.1 - 85.3 | 2,148 | 1691 ( 78.7 ) | 76.9 - 80.4 | 2,148 | 778 ( 36.2 ) | 34.2 - 38.3 | 2,148 | 105 ( 4.9 ) | 4.1 - 5.9 |
|  | 30-34 | 1,937 | 1658 ( 85.6 ) | 84.0 - 87.1 | 1,937 | 1566 ( 80.8 ) | 79.0 - 82.5 | 1,937 | 764 ( 39.4 ) | 37.3 - 41.6 | 1,937 | 109 ( 5.6 ) | 4.7 - 6.7 |
|  | 35-39 | 1,852 | 1584 ( 85.5 ) | 83.9 - 87.1 | 1,851 | 1504 ( 81.3 ) | 79.4 - 83.0 | 1,852 | 763 ( 41.2 ) | 39.0 - 43.5 | 1,852 | 105 ( 5.7 ) | 4.7 - 6.8 |
|  | 40-44 | 1,389 | 1217 ( 87.6 ) | 85.8 - 89.2 | 1,389 | 1145 ( 82.4 ) | 80.3 - 84.3 | 1,388 | 573 ( 41.3 ) | 38.7 - 43.9 | 1,389 | 103 ( 7.4 ) | 6.2 - 8.9 |
|  | 45-49 | 881 | 792 ( 89.9 ) | 87.7 - 91.7 | 882 | 734 ( 83.2 ) | 80.6 - 85.5 | 882 | 415 ( 47.1 ) | 43.8 - 50.4 | 882 | 63 ( 7.1 ) | 5.6 – 9.0 |
| Education | Basic | 4,084 | 3394 ( 83.1 ) | 81.9 - 84.2 | 4,084 | 3173 ( 77.7 ) | 76.4 - 78.9 | 4,083 | 1545 ( 37.8 ) | 36.4 - 39.3 | 4,084 | 246 ( 6 ) | 5.3 - 6.8 |
|  | Higher | 623 | 570 ( 91.5 ) | 89.0 - 93.4 | 623 | 551 ( 88.4 ) | 85.7 - 90.7 | 623 | 329 ( 52.8 ) | 48.9 - 56.7 | 624 | 19 ( 3 ) | 2.0 - 4.7 |
|  | No education | 3,036 | 2575 ( 84.8 ) | 83.5 – 86.0 | 3,037 | 2419 ( 79.7 ) | 78.2 – 81.0 | 3,037 | 976 ( 32.1 ) | 30.5 - 33.8 | 3,037 | 244 ( 8 ) | 7.1 - 9.1 |
|  | Secondary | 5,322 | 4546 ( 85.4 ) | 84.4 - 86.3 | 5,322 | 4287 ( 80.6 ) | 79.5 - 81.6 | 5,321 | 2312 ( 43.5 ) | 42.1 - 44.8 | 5,321 | 297 ( 5.6 ) | 5.0 - 6.2 |
| Province | Bagmati | 2,763 | 2183 ( 79.0 ) | 77.5 - 80.5 | 2,763 | 2078 ( 75.2 ) | 73.6 - 76.8 | 2,762 | 1009 ( 36.5 ) | 34.8 - 38.3 | 2,763 | 51 ( 1.8 ) | 1.4 - 2.4 |
|  | Gandaki | 1,292 | 1037 ( 80.3 ) | 78.0 - 82.3 | 1,292 | 953 ( 73.8 ) | 71.3 - 76.1 | 1,292 | 514 ( 39.8 ) | 37.1 - 42.5 | 1,292 | 46 ( 3.6 ) | 2.7 - 4.7 |
|  | Karnali | 781 | 716 ( 91.7 ) | 89.5 - 93.4 | 782 | 650 ( 83.1 ) | 80.3 - 85.6 | 781 | 539 ( 69.0 ) | 65.7 - 72.2 | 782 | 204 ( 26.1 ) | 23.1 - 29.3 |
|  | Koshi | 2,193 | 1895 ( 86.4 ) | 84.9 - 87.8 | 2,193 | 1825 ( 83.2 ) | 81.6 - 84.7 | 2,193 | 912 ( 41.6 ) | 39.5 - 43.7 | 2,193 | 72 ( 3.3 ) | 2.6 - 4.1 |
|  | Lumbini | 2,457 | 2065 ( 84.0 ) | 82.5 - 85.4 | 2,456 | 1949 ( 79.4 ) | 77.7 - 80.9 | 2,457 | 985 ( 40.1 ) | 38.2 – 42.0 | 2,457 | 99 ( 4 ) | 3.3 - 4.9 |
|  | Madhesh | 2,466 | 2126 ( 86.2 ) | 84.8 - 87.5 | 2,467 | 2101 ( 85.2 ) | 83.7 - 86.5 | 2,467 | 395 ( 16.0 ) | 14.6 - 17.5 | 2,466 | 116 ( 4.7 ) | 3.9 - 5.6 |
|  | Sudurpashchim | 1,113 | 1064 ( 95.6 ) | 94.2 - 96.7 | 1,114 | 875 ( 78.5 ) | 76.0 - 80.9 | 1,113 | 808 ( 72.6 ) | 69.9 - 75.1 | 1,114 | 218 ( 19.6 ) | 17.3 - 22 |
| Ethnicity | Brahmin/Chhetri | 3,735 | 3588 ( 96.1 ) | 95.4 - 96.6 | 3,735 | 3253 ( 87.1 ) | 86.0 - 88.1 | 3,735 | 2847 ( 76.2 ) | 74.8 - 77.6 | 3,735 | 457 ( 12.2 ) | 11.2 - 13.3 |
|  | Dalit | 1,933 | 1587 ( 82.1 ) | 80.3 - 83.7 | 1,934 | 1448 ( 74.9 ) | 72.9 - 76.8 | 1,934 | 761 ( 39.3 ) | 37.2 - 41.5 | 1,934 | 160 ( 8.3 ) | 7.1 - 9.6 |
|  | Janajati | 4,879 | 3709 ( 76.0 ) | 74.8 - 77.2 | 4,878 | 3562 ( 73.0 ) | 71.8 - 74.2 | 4,879 | 974 ( 20.0 ) | 18.9 - 21.1 | 4,878 | 70 ( 1.4 ) | 1.1 - 1.8 |
|  | Madhesi | 1,942 | 1778 ( 91.6 ) | 90.2 - 92.7 | 1,943 | 1756 ( 90.4 ) | 89.0 - 91.6 | 1,942 | 526 ( 27.1 ) | 25.2 - 29.1 | 1,942 | 117 ( 6 ) | 5.1 - 7.2 |
|  | Muslim | 562 | 411 ( 73.1 ) | 69.3 - 76.6 | 562 | 399 ( 71.0 ) | 67.1 - 74.6 | 562 | 47 ( 8.4 ) | 6.3 - 10.9 | 562 | 0 ( 0 ) | 0 - 0.7 |
|  | Other | 14 | 12 ( 85.7 ) | 60.1 – 96.0 | 14 | 12 ( 85.7 ) | 60.1 – 96.0 | 14 | 8 ( 57.1 ) | 32.6 - 78.6 | 14 | 2 ( 14.3 ) | 4.0 - 39.9 |
| Residence | Rural | 4,081 | 3391 ( 83.1 ) | 81.9 - 84.2 | 4,081 | 3126 ( 76.6 ) | 75.3 - 77.9 | 4,081 | 1672 ( 41.0 ) | 39.5 - 42.5 | 4,081 | 373 ( 9.1 ) | 8.3 - 10.1 |
|  | Urban | 8,985 | 7695 ( 85.6 ) | 84.9 - 86.4 | 8,985 | 7304 ( 81.3 ) | 80.5 - 82.1 | 8,984 | 3491 ( 38.9 ) | 37.9 - 39.9 | 8,985 | 433 ( 4.8 ) | 4.4 - 5.3 |
| Gender of Household head | Female | 4,568 | 3811 ( 83.4 ) | 82.3 - 84.5 | 4,568 | 3593 ( 78.7 ) | 77.4 - 79.8 | 4,568 | 1647 ( 36.1 ) | 34.7 - 37.5 | 4,568 | 302 ( 6.6 ) | 5.9 - 7.4 |
|  | Male | 8,497 | 7274 ( 85.6 ) | 84.8 - 86.3 | 8,497 | 6836 ( 80.5 ) | 79.6 - 81.3 | 8,497 | 3516 ( 41.4 ) | 40.3 - 42.4 | 8,497 | 504 ( 5.9 ) | 5.4 - 6.5 |
| Age of household head | 15-24 | 615 | 492 ( 80.0 ) | 76.7 – 83.0 | 616 | 465 ( 75.5 ) | 71.9 - 78.7 | 616 | 167 ( 27.1 ) | 23.7 - 30.8 | 616 | 34 ( 5.5 ) | 4.0 - 7.6 |
|  | 25-34 | 2,288 | 1898 ( 83.0 ) | 81.4 - 84.4 | 2,288 | 1795 ( 78.5 ) | 76.7 - 80.1 | 2,289 | 724 ( 31.6 ) | 29.8 - 33.6 | 2,288 | 124 ( 5.4 ) | 4.6 - 6.4 |
|  | 35-44 | 3,938 | 3299 ( 83.8 ) | 82.6 - 84.9 | 3,937 | 3120 ( 79.2 ) | 78.0 - 80.5 | 3,938 | 1528 ( 38.8 ) | 37.3 - 40.3 | 3,938 | 256 ( 6.5 ) | 5.8 - 7.3 |
|  | 45-54 | 3,161 | 2711 ( 85.8 ) | 84.5 - 86.9 | 3,161 | 2559 ( 81.0 ) | 79.5 - 82.3 | 3,161 | 1289 ( 40.8 ) | 39.1 - 42.5 | 3,161 | 192 ( 6.1 ) | 5.3 – 7.0 |
|  | 55-64 | 1,773 | 1550 ( 87.4 ) | 85.8 - 88.9 | 1,773 | 1450 ( 81.8 ) | 79.9 - 83.5 | 1,773 | 797 ( 45.0 ) | 42.6 - 47.3 | 1,773 | 105 ( 5.9 ) | 4.9 - 7.1 |
|  | Over 64 | 1,290 | 1135 ( 88.0 ) | 86.1 - 89.6 | 1,290 | 1039 ( 80.5 ) | 78.3 - 82.6 | 1,290 | 659 ( 51.1 ) | 48.4 - 53.8 | 1,290 | 95 ( 7.4 ) | 6.1 - 8.9 |
| W | Poorest | 2,280 | 1869 ( 82.0 ) | 80.3 - 83.5 | 2,281 | 1615 ( 70.8 ) | 68.9 - 72.6 | 2,281 | 1152 ( 50.5 ) | 48.5 - 52.6 | 2,281 | 331 ( 14.5 ) | 13.1 – 16.0 |
| Characteristic | Characteristic | N | Prevalence<br>n (%) | (95% CI) | N | Prevalence<br>n (%) | (95% CI) | N | Prevalence<br>n (%) | (95% CI) | N | Prevalence<br>n (%) | (95% CI) |
| ealth index | Poorer | 2,443 | 2077 ( 85.0 ) | 83.5 - 86.4 | 2,443 | 1953 ( 79.9 ) | 78.3 - 81.5 | 2,442 | 874 ( 35.8 ) | 33.9 - 37.7 | 2,443 | 155 ( 6.3 ) | 5.4 - 7.4 |
|  | Middle | 2,636 | 2213 ( 84.0 ) | 82.5 - 85.3 | 2,636 | 2121 ( 80.5 ) | 78.9 - 81.9 | 2,636 | 882 ( 33.5 ) | 31.7 - 35.3 | 2,636 | 151 ( 5.7 ) | 4.9 - 6.7 |
|  | Richer | 2,868 | 2449 ( 85.4 ) | 84.1 - 86.6 | 2,868 | 2343 ( 81.7 ) | 80.2 - 83.1 | 2,868 | 1004 ( 35 ) | 33.3 - 36.8 | 2,868 | 118 ( 4.1 ) | 3.4 - 4.9 |
|  | Richest | 2,839 | 2478 ( 87.3 ) | 86.0 - 88.5 | 2,839 | 2398 ( 84.5 ) | 83.1 - 85.8 | 2,840 | 1251 ( 44 ) | 42.2 - 45.9 | 2,839 | 52 ( 1.8 ) | 1.4 - 2.4 |
| Total |  | 13,065 | 11085 ( 84.8 ) | 84.2 - 85.4 | 13,065 | 10429 ( 79.8 ) | 79.1 - 80.5 | 13,065 | 5163 ( 39.5 ) | 38.7 - 40.4 | 13,065 | 806 ( 6.2 ) | 5.8 - 6.6 |

### Study variables

Our study sought to describe menstrual discriminatory practices among Nepalese women. These practices include restrictions on activities such as entering the temple, participating in religious activities, cooking, handling food items, fetching water, eating with the family, and the change in sleeping arrangements within the household during menstruation. In our analyses, menstrual restriction activities were grouped into four binary composite outcomes: a) any form of menstrual restriction defined as any menstrual related exclusions, b) religious restrictions defined as exclusions from entering the temple or participating in religious activities, c) household level restriction was defined as exclusions from fetching water, cooking and handling food items, eating with the family, sleeping in the main house, and sleeping with the husband d) *Chhaupadi* defined as exclusion from sleeping in the main house as described previously (ICF, 2023). For each composite binary outcome, {1} indicate an experience of at least one restriction within the defined sub-group and {0} otherwise. Variations in menstrual restrictions was assessed in the regression models using the participants characteristics (age, ethnicity/caste, education), household characteristics (age and sex of the household head, and wealth) and regional characteristics (urbanicity and region (province) of residence). Detailed description of these variables has been reported previously (ICF, 2023).

### Statistical analyses

The prevalence of menstrual restrictions by social demographic characteristics were estimated accounting for the study design and used the Taylor linearization approach to account for the complex sampling design and clustering.

### Spatial Analyses

The risk maps were created using an adaptive bandwidth in a weighted Gaussian kernel density estimation methodology (Larmarange et al., 2011) to yield spatially continuous maps and interpolate georeferenced outcomes of menstrual restriction.

### Local cluster detection

We applied the Bernoulli probability model within SaTScan software to identify geographic areas with significantly higher menstrual restriction cases than expected by chance. We specified a circular, non-overlapping scanning window that systematically moves across the geographical space, with the radius varying from zero to a maximum radius determined based on a predetermined maximum enclosed population size (up to 50% in these analyses). For each location and window size, the software tallies the observed cases while the expected cases are computed assuming a uniform distribution across the population. The scan statistic compares case counts inside and outside each circle, generating an observed log likelihood statistic. To test the null hypothesis of complete spatial randomness, Monte-Carlo simulation with 999 replications is used to created permutations of the observed cases across all data locations which is used to generate the simulated log likelihoods. The observed log likelihood and the simulated log likelihoods are the used to calculate the p-value for statistical significance testing. The Gini coefficient determined optimal non-overlapping cluster sizes, and clusters with a p-value <0.05 were considered significant. No covariate adjustments were made in either the weighted Gaussian kernel density estimation or the local cluster analyses. This is because our focus was to describe spatial variations in menstrual restrictions without accounting for specific causes.

### Regression analyses

We used a weighted modified Poisson regression model (weighted to accommodate the NDHS sampling design weights) to assess the association between menstrual restriction outcomes and the socioeconomic and demographic factors. To assess the extent to which variations on menstrual stigma are accounted for by unmeasured factor variable at the regional level, we incorporated region (province) as a fixed effect in the regression model. The McFadden’s pseudo R^2^ was used to quantify the contribution of each socioeconomic and demographic variable in the regression models. Statistical analyses were performed in software R version 4.3.2, local clusters were detected using SaTScan version 10.1.2 and maps were produced using QGIS version 3.34.2. All analyses accounted for the survey design.

## Results

Of 14,845 women aged 15 to 49 years who participated in the 2022 NDHS, 1780 (12%) had incomplete data on menstrual restrictions. Therefore, our analysis included 13,065 (88%) study participants who reported at least one menstrual event within the last one year preceding the survey. The distribution of the study participants by social-demographic characteristics are presented in **(Supp. Table 1)**

### Prevalence of menstrual restrictions

The overall prevalence of experiencing any type of menstrual restriction in Nepal was 84.8 % (95% CI: 84.2-85.4). Among the seven provinces, Sudurpashchim had the highest prevalence of experiencing any form of menstrual restrictions (95.6% [95% CI: 94.2-96.7]) followed by Karnali (91.7% [95%CI: 89.5-93.4]) whist Bagmati reported the lowest prevalence (79.0% [95% CI: 77.5-80.5]). Furthermore, the prevalence of experiencing any form of menstrual restriction was highest among older women (89.9% [95%CI: 87.7 - 91.7] among women aged 45-49), those with higher education (91.5% [95%CI: 89.0 – 93.4]), Brahmin/Chhetri ethnic background (96.1% [95%CI: 95.4-96.6]), residing in households led by men (85.6% [95%CI: 84.8-86.3]) or led by older household heads (88.0% [95%CI: 86.1-89.6] among household heads older than 64 years) and those residing in wealthy households (87.3% [95%CI: 86.0-88.5]). However, because of the high prevalence of experiencing any form of menstrual restrictions in this setting, variations within socioeconomic and demographic factors were moderate to small (**Table 1).** Among the other forms of menstrual discriminatory practices, religious restrictions were the most prevalent (79.8% [95%CI: 79.1–80.5]), followed by household level restrictions (39.5% [95%CI: 38.7–40.4]) and *Chhaupadi* (6.2% [95%CI: 5.8–6.6]) in that order. Further description of the socioeconomic and demographic factors for each form of menstrual restriction is presented in **Table 1**.

### Spatial Heterogeneity of Menstrual Restrictions

At the region (province) level, there was moderate variations in the prevalence of any menstrual restriction (range: 79.0% to 95.6%) and religious restrictions (range: 73.8% to 85.2%). However, we observed marked regional variations in household level restrictions (range: 16.0% to 76.2%), and *Chhaupadi* (range: 1.8% to 26.1%). Geospatial analysis using the adaptive Gaussian kernel density methodology revealed clear spatial variation in prevalence of menstrual restrictions across the study area. The kernel-derived prevalence of experiencing any type of menstrual restriction varied between 23.7% and 100%, and with mild variation defined by the country’s topography (**Figure 2**A). Similarly, the high prevalence of religious restrictions was geographically restricted to the densely populated *Terai* and Hilly areas (**Figure 2**B). Local cluster analyses of religious restrictions revealed 17 statistically significant high-prevalence clusters (p<0.05), however, the ratio between the risk inside and outside the clusters (RR range: 1.13 and 1.27) were small (**Table**) suggesting a near homogeneous distribution in prevalence across the country.

**Figure 1.**
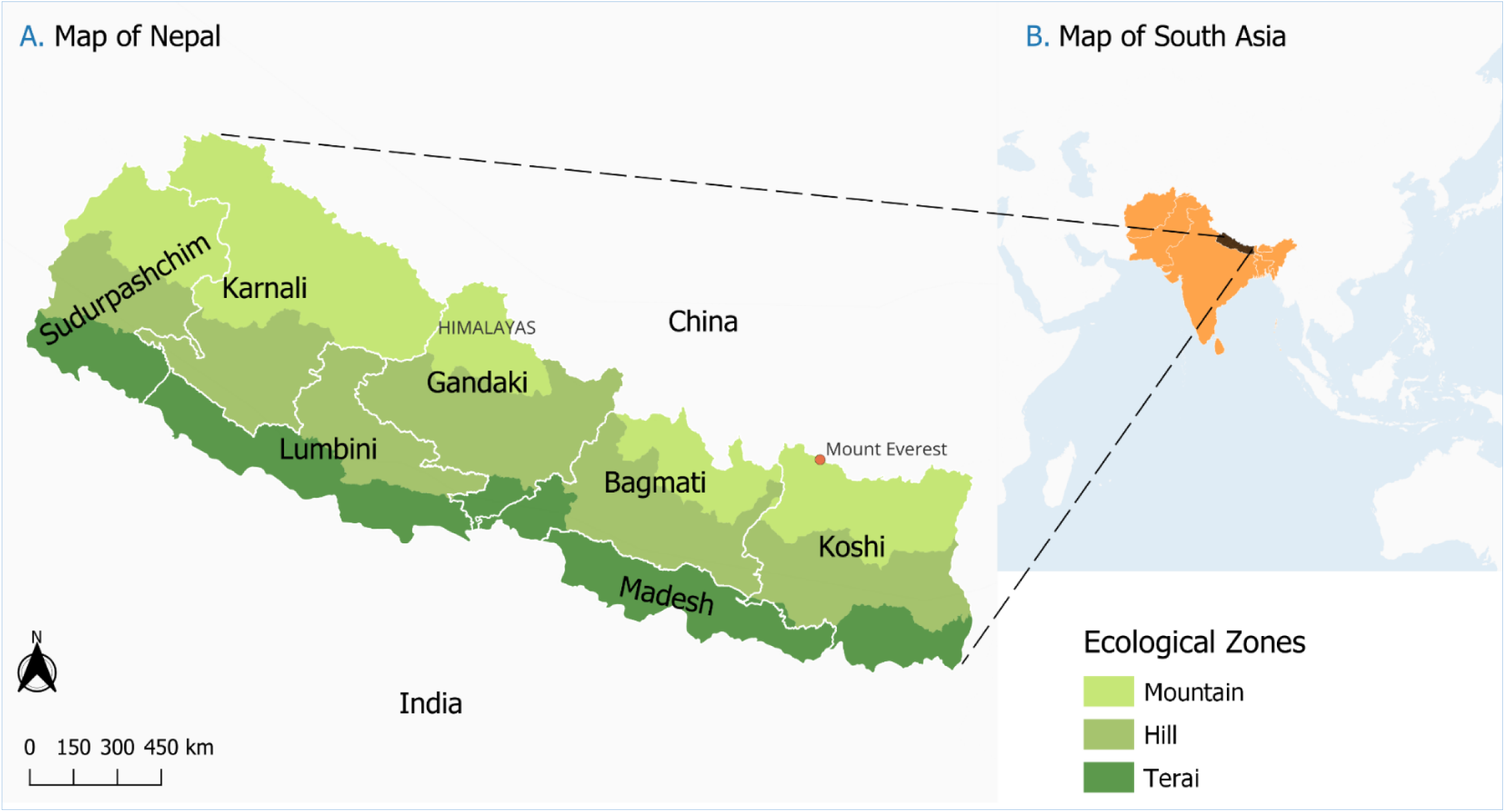
Map of the study area showing the various ecological zones, first level administrative divisions and the neighboring countries.

**Figure 2.**
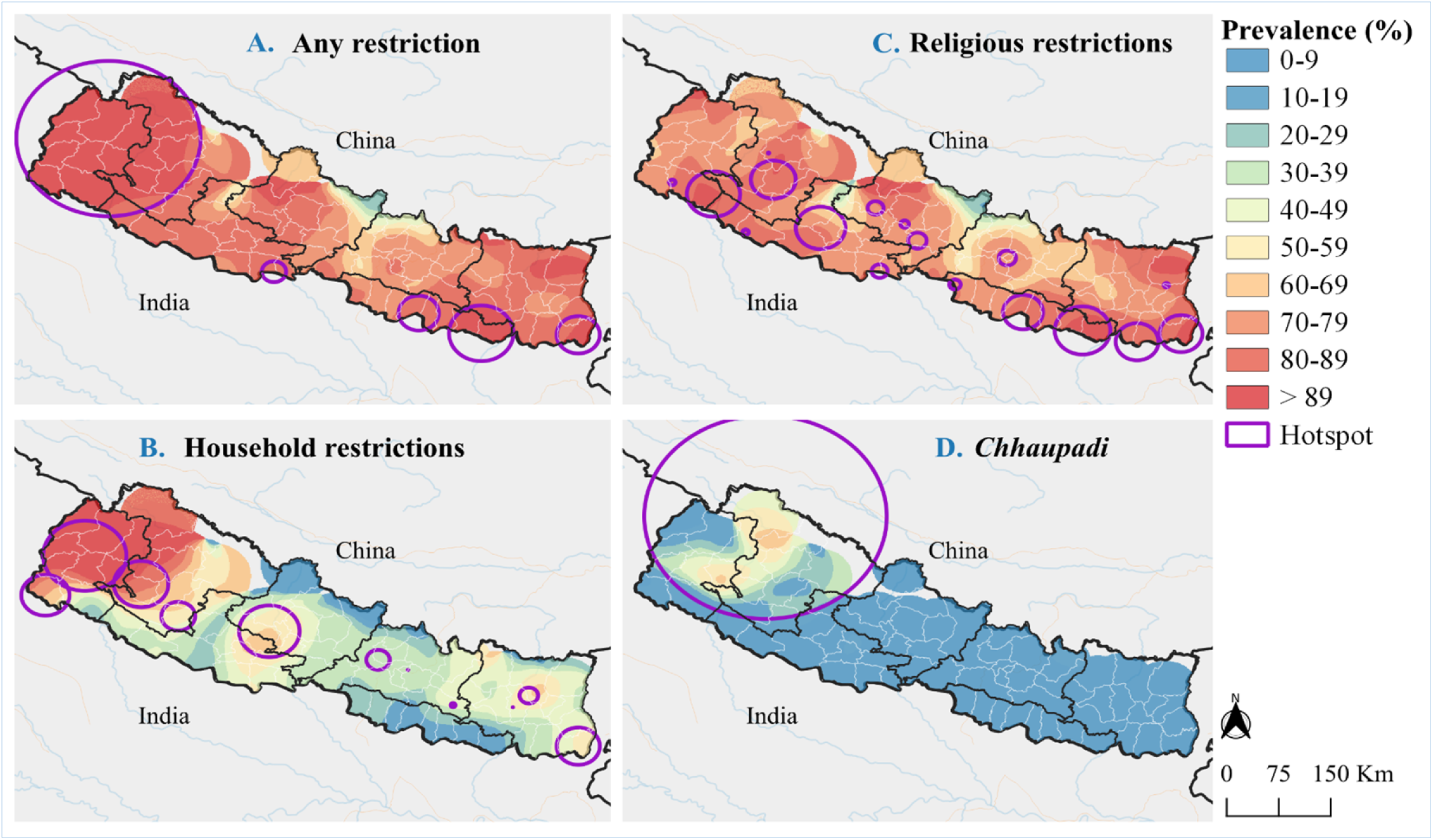
Continuous surface maps of prevalence and clusters of high prevalence of menstrual discriminatory practices in Nepal. Panels (**A**) to (**D**) show the spatial variations in the prevalence of the various forms of menstrual restrictions and the corresponding clusters of high prevalence.

The kernel-derived prevalence of menstrual restriction within household varied markedly over the geographical area, ranging between 0% and 100% and between 0% and 66.9% for *Chhaupadi*. These analyses reveal a clear region of high prevalence (middle and western region of Nepal) and low prevalence in the eastern region of Nepal (**Figure 2**). Local cluster analyses of household level restrictions and *Chhaupadi* revealed 11 and 1 statistically significant clusters respectively. The prevalence ratios for the detected high-prevalence clusters ranged from 1.32 to 2.34 for household level restrictions, and 7.45 for *Chhaupadi*. In all the local cluster analyses, the primary clusters were located in either Sudurpashchim or Karnali or both (**Figure 2**, **Table**).

### Predictors of menstrual restrictions

In the multivariable analysis, older women aged between 35-39, 40-44, and 45-49 were associated with a 5% (aPR=1.05 [95%CI: 1.01–1.09], *P*=0.021), 5% (aPR=1.05 [95%CI: 1.01–1.10], *P* =0.011), and 6% (aPR=1.06 [95%CI: 1.01–1.11], *P* =0.022) increase in the likelihood of experiencing restrictions on religious activities respectively. However, there were no clear association with household level restrictions or *Chhaupadi*. Compared to women with no education, women with primary, secondary and higher education were associated with a 13% (aPR=1.13 [95%CI: 1.05–1.22], *P*=0.002), 10% (aPR=1.10 [95%CI: 1.00–1.21], *P* =0.041), and 17% (aPR=1.17[95%CI: 1.03–1.34], *P* =0.015) increase in the likelihood of experiencing household level restrictions, however, there was no evidence of an associations between education and the other forms of menstrual restrictions. Ethnicity/caste was associated with variations in all forms of menstrual restrictions whilst rural/urban residence was only significantly associated with *Chhaupadi* in the univariable models but not in the multivariable models (**Supp. Table 2**). Women residing in households led by a female head were associated with 11% (aPR=0.89 [95%CI: 0.84–0.94], p<0.001) lower likelihood of experiencing household level restrictions but no evidence of an association with restrictions on religious activities or *Chhaupadi* (**Table 2**). In addition, older household heads were associated with an increased risk of experiencing household level menstrual restrictions and *Chhaupadi*. On one hand, increased household’s wealth was associated with an increased risk of experiencing restrictions on religious activities. On the other hand, increased household’s wealth was significantly associated with protection against *Chhaupadi* (**Table 2**).

**Table 2:** Multivariable Poisson regression of menstrual restrictions by social-demographic and economic factors.

|  | Model1: Any restriction |  |  | Model2: Religious restrictions |  |  | Model3: Household level restrictions |  |  | Model4: *Chhaupadi* |  |  |
| --- | --- | --- | --- | --- | --- | --- | --- | --- | --- | --- | --- | --- |
| Characteristic | aPR | 95% CI | P-value | aPR | 95% CI | P-value | aPR | 95% CI | P-value | aPR | 95% CI | P-value |
| **Age (ref: 15-19 years)** |  |  |  |  |  |  |  |  |  |  |  |  |
| 20-24 | 1.01 | 0.98-1.03 | 0.723 | 1.00 | 0.97-1.04 | 0.852 | 0.94 | 0.89-1.01 | 0.090 | 0.93 | 0.76-1.13 | 0.450 |
| 25-29 | 1.01 | 0.98-1.04 | 0.601 | 1.00 | 0.97-1.04 | 0.830 | 0.95 | 0.88-1.03 | 0.246 | 0.83 | 0.65-1.07 | 0.154 |
| 30-34 | 1.03 | 1.00-1.07 | 0.064 | 1.03 | 0.99-1.07 | 0.115 | 1.03 | 0.95-1.12 | 0.463 | 0.93 | 0.73-1.19 | 0.567 |
| 35-39 | 1.04 | 1.01-1.08 | 0.024 | 1.05 | 1.01-1.09 | 0.021 | 1.03 | 0.95-1.12 | 0.483 | 0.87 | 0.67-1.12 | 0.285 |
| 40-44 | 1.06 | 1.02-1.10 | 0.001 | 1.05 | 1.01-1.10 | 0.011 | 1.02 | 0.94-1.11 | 0.643 | 1.12 | 0.82-1.53 | 0.484 |
| 45-49 | 1.07 | 1.03-1.12 | 0.001 | 1.06 | 1.01-1.11 | 0.022 | 1.09 | 0.99-1.21 | 0.070 | 1.05 | 0.76-1.45 | 0.754 |
| **Highest level of education (ref: No education)** |  |  |  |  |  |  |  |  |  |  |  |  |
| Basic | 0.99 | 0.96-1.02 | 0.599 | 0.98 | 0.95-1.01 | 0.220 | 1.13 | 1.05-1.22 | 0.002 | 0.94 | 0.76-1.17 | 0.600 |
| Secondary | 0.99 | 0.96-1.02 | 0.642 | 0.98 | 0.95-1.02 | 0.339 | 1.10 | 1.00-1.21 | 0.041 | 0.92 | 0.72-1.18 | 0.529 |
| Higher | 1.03 | 0.98-1.08 | 0.224 | 1.03 | 0.98-1.09 | 0.192 | 1.17 | 1.03-1.34 | 0.015 | 0.95 | 0.55-1.65 | 0.863 |
| **Residence (ref: Rural)** |  |  |  |  |  |  |  |  |  |  |  |  |
| Urban | 1.02 | 0.98-1.05 | 0.37 | 1.02 | 0.98-1.06 | 0.378 | 1.02 | 0.93-1.11 | 0.707 | 0.81 | 0.63-1.05 | 0.11 |
| **Caste/Ethnicity (ref: Janajati)** |  |  |  |  |  |  |  |  |  |  |  |  |
| Brahmin/Chhetri | 1.24 | 1.20-1.28 | <0.001 | 1.19 | 1.14-1.23 | <0.001 | 3.53 | 3.15-3.95 | <0.001 | 5.41 | 3.76-7.78 | <0.001 |
| Dalit | 1.07 | 1.03-1.12 | 0.002 | 1.02 | 0.97-1.08 | 0.376 | 2.26 | 1.97-2.59 | <0.001 | 4.00 | 2.70-5.92 | <0.001 |
| Madhesi | 1.17 | 1.12-1.22 | <0.001 | 1.15 | 1.09-1.20 | <0.001 | 2.08 | 1.68-2.58 | <0.001 | 4.75 | 2.61-8.63 | <0.001 |
| Muslim | 0.93 | 0.81-1.07 | 0.299 | 0.90 | 0.78-1.04 | 0.157 | 0.64 | 0.42-0.99 | 0.047 | 0.00 | 0.00-0.00 | <0.001 |
| Other | 1.12 | 0.85-1.48 | 0.420 | 1.13 | 0.85-1.49 | 0.397 | 3.06 | 1.62-5.75 | 0.001 | 13.57 | 1.98-92.98 | 0.008 |
| **Region/Province (ref: Bagmati)** |  |  |  |  |  |  |  |  |  |  |  |  |
| Gandaki | 1.04 | 0.96-1.12 | 0.325 | 1.01 | 0.93-1.11 | 0.791 | 1.13 | 0.98-1.32 | 0.103 | 1.57 | 0.93-2.64 | 0.094 |
| Karnali | 1.14 | 1.08-1.22 | <0.001 | 1.17 | 1.09-1.25 | <0.001 | 1.36 | 1.22-1.53 | <0.001 | 5.19 | 3.29-8.18 | <0.001 |
| Koshi | 1.13 | 1.06-1.20 | <0.001 | 1.16 | 1.09-1.24 | <0.001 | 1.22 | 1.07-1.40 | 0.004 | 1.34 | 0.75-2.39 | 0.329 |
| Characteristic | aPR | 95% CI | P-value | aPR | 95% CI | P-value | aPR | 95% CI | P-value | aPR | 95% CI | P-value |
| Lumbini | 1.08 | 1.02-1.14 | 0.014 | 1.08 | 1.02-1.16 | 0.016 | 1.06 | 0.93-1.22 | 0.370 | 1.46 | 0.81-2.62 | 0.206 |
| Madhesh | 1.11 | 1.03-1.19 | 0.005 | 1.16 | 1.08-1.26 | <0.001 | 0.47 | 0.36-0.61 | <0.001 | 1.40 | 0.72-2.70 | 0.319 |
| Sudurpashchim | 1.19 | 1.13-1.26 | <0.001 | 1.07 | 1.00-1.16 | 0.063 | 1.54 | 1.38-1.72 | <0.001 | 4.70 | 2.92-7.57 | <0.001 |
| **Gender of the household head (ref: Male)** |  |  |  |  |  |  |  |  |  |  |  |  |
| Female | 0.99 | 0.97-1.01 | 0.274 | 1.00 | 0.97-1.02 | 0.792 | 0.89 | 0.84-0.94 | <0.001 | 1.04 | 0.89-1.2 | 0.646 |
| **Age of the household head (ref 15-24 years)** |  |  |  |  |  |  |  |  |  |  |  |  |
| 25-34 | 1.03 | 0.97-1.09 | 0.412 | 1.02 | 0.96-1.09 | 0.545 | 1.24 | 1.06-1.46 | 0.008 | 1.19 | 0.85-1.67 | 0.310 |
| 35-44 | 1.02 | 0.96-1.08 | 0.603 | 1.00 | 0.94-1.07 | 0.919 | 1.55 | 1.34-1.79 | <0.001 | 1.47 | 1.05-2.06 | 0.024 |
| 45-54 | 1.04 | 0.98-1.10 | 0.191 | 1.03 | 0.96-1.10 | 0.394 | 1.62 | 1.39-1.89 | <0.001 | 1.52 | 1.08-2.14 | 0.018 |
| 55-64 | 1.06 | 1.00-1.13 | 0.046 | 1.04 | 0.97-1.11 | 0.266 | 1.78 | 1.52-2.07 | <0.001 | 1.48 | 1.05-2.08 | 0.025 |
| Over 64 | 1.06 | 1.00-1.13 | 0.049 | 1.02 | 0.95-1.09 | 0.659 | 1.97 | 1.69-2.29 | <0.001 | 1.89 | 1.31-2.74 | 0.001 |
| **Wealth Index (ref: Poorest)** |  |  |  |  |  |  |  |  |  |  |  |  |
| Poorer | 1.07 | 1.03-1.12 | 0.001 | 1.15 | 1.09-1.21 | <0.001 | 0.96 | 0.89-1.04 | 0.297 | 0.79 | 0.62-1.01 | 0.058 |
| Middle | 1.06 | 1.01-1.12 | 0.012 | 1.15 | 1.09-1.22 | <0.001 | 0.92 | 0.83-1.02 | 0.110 | 0.81 | 0.62-1.07 | 0.144 |
| Richer | 1.09 | 1.03-1.14 | 0.001 | 1.18 | 1.11-1.25 | <0.001 | 0.91 | 0.82-1.01 | 0.078 | 0.58 | 0.43-0.78 | <0.001 |
| Richest | 1.11 | 1.05-1.18 | <0.001 | 1.23 | 1.16-1.32 | <0.001 | 0.90 | 0.80-1.02 | 0.091 | 0.26 | 0.17-0.39 | <0.001 |

**Table 3:** Characteristics of clusters of high prevalence.

| Restriction | Hotspot | Prevalence Risk | P-Value | Radius (KM) |
| --- | --- | --- | --- | --- |
| Any restriction | 1 | 1.08 | 0.003 | 28.68 |
|  | 2 | 1.11 | <0.001 | 30.79 |
|  | 3 | 1.13 | <0.001 | 45.95 |
|  | 4 | 1.13 | 0.013 | 17.23 |
|  | 5 | 1.16 | <0.001 | 127.88 |
| Religious restrictions | 1 | 1.12 | 0.001 | 12.40 |
|  | 2 | 1.13 | 0.02 | 35.18 |
|  | 3 | 1.14 | <0.001 | 30.89 |
|  | 4 | 1.16 | <0.001 | 28.68 |
|  | 5 | 1.18 | <0.001 | 31.67 |
|  | 6 | 1.18 | 0.014 | 6.07 |
|  | 7 | 1.19 | <0.001 | 37.65 |
|  | 8 | 1.20 | <0.001 | 30.79 |
|  | 9 | 1.21 | 0.042 | 4.62 |
|  | 10 | 1.22 | <0.001 | 40.03 |
|  | 11 | 1.24 | <0.001 | 11.32 |
|  | 12 | 1.24 | 0.003 | 13.28 |
|  | 13 | 1.25 | 0.019 | 8.22 |
|  | 14 | 1.27 | <0.001 | 0.89 |
|  | 15 | 1.27 | 0.003 | 3.69 |
|  | 16 | 1.27 | 0.005 | 11.26 |
|  | 17 | 1.27 | 0.024 | 4.57 |
| Household restrictions | 1 | 1.32 | 0.001 | 30.79 |
|  | 2 | 1.41 | 0.001 | 42.58 |
|  | 3 | 1.57 | 0.001 | 33.67 |
|  | 4 | 1.59 | 0.001 | 16.45 |
|  | 5 | 1.69 | 0.001 | 23.68 |
|  | 6 | 1.77 | 0.001 | 3.41 |
|  | 7 | 1.83 | 0.001 | 13.14 |
|  | 8 | 1.86 | 0.024 | 0.00 |
|  | 9 | 1.91 | 0.001 | 38.12 |
|  | 10 | 2.14 | 0.001 | 0.00 |
|  | 11 | 2.34 | 0.001 | 57.79 |
| *Chhaupadi* | 1 | 7.45 | <0.001 | 169.05 |

The final multivariable regression model accounted for 4%, 3%, 24%, and 23% of the variability in any form of menstrual restriction, restriction on religious activities, household level restrictions and *Chhaupadi* respectively. However, the variability accounted for by the variable ethnicity/caste was 3% for any form of menstrual restriction, 2% for restriction on religious activities, 19% for household level restrictions and 11% for *Chhaupadi*.

## Discussion

In the global pursuit of improved Water, Sanitation, and Hygiene (WASH), menstrual health and hygiene (MHH) persist as pressing challenges in low and middle-income countries (LMICs) directly contributing to socio-economic inequity. Although there has been significant progress in other areas of WASH, millions of women still encounter obstacles in managing their menstrual periods safely and with dignity (Crawford et al., 2014; Health, 2018; Health–Americas, 2022). Our results indicate that Nepal continues to bear a disproportionate burden of menstrual-related discrimination, which manifest as exclusions from participating in social and religious activities. These findings are consistent with recent studies, which showed that *Chhaupadi* is most prevalent among the Brahmin/Chhetri ethnic community in the mid-western part of Nepal (Amery et al., 2023; Thakuri et al., 2021; UNFPA, 2019; UNICEF, 2020, 2022).

Overall, 84.8% of women in Nepal experienced some form of menstrual restriction that were characterized by mild geographical variations, likely due to the high prevalence observed across the entire country. Among the various forms of menstrual discriminatory practices considered, restrictions on religious activities were the most prevalent (79.8%) nationwide. This estimate reflects a 13% decline from the 2019 figures (UNICEF, 2020), a trend that could be attributed to sustained efforts various government and non-governmental agencies to eradicate menstrual discrimination (Secretariat & Durbar, 2015; ActionAid, 2024). The prevalence of *Chhaupadi* was highest in the mid and western part of Nepal, highlighting the role of regional, ethnic and religious dynamics in shaping menstrual discriminatory practices. Our estimates in Karnali and Sudurpashchim provinces are similar with the figures in the United Nations Urgency report (UNICEF, 2020), reflecting minimal gains in the efforts to eradicate the harmful practices. Achham and Deilekh districts were associated with the highest *Chhaupadi* case count per square kilometre (**Figure 3B**). This is noteworthy, considering that the two districts have previously been targeted with interventions that include destruction of the menstrual huts and prosecution of those practising the vice (Adhikari, 2020; Gurung, 2017; Vaughn, 2019). Our findings are consistent with prior studies that cite gaps in the enforcement of legislative interventions aimed at curbing the practice (Baumann, Merante, et al., 2021; Gurung, 2017; Vaughn, 2019).

**Figure 3.**
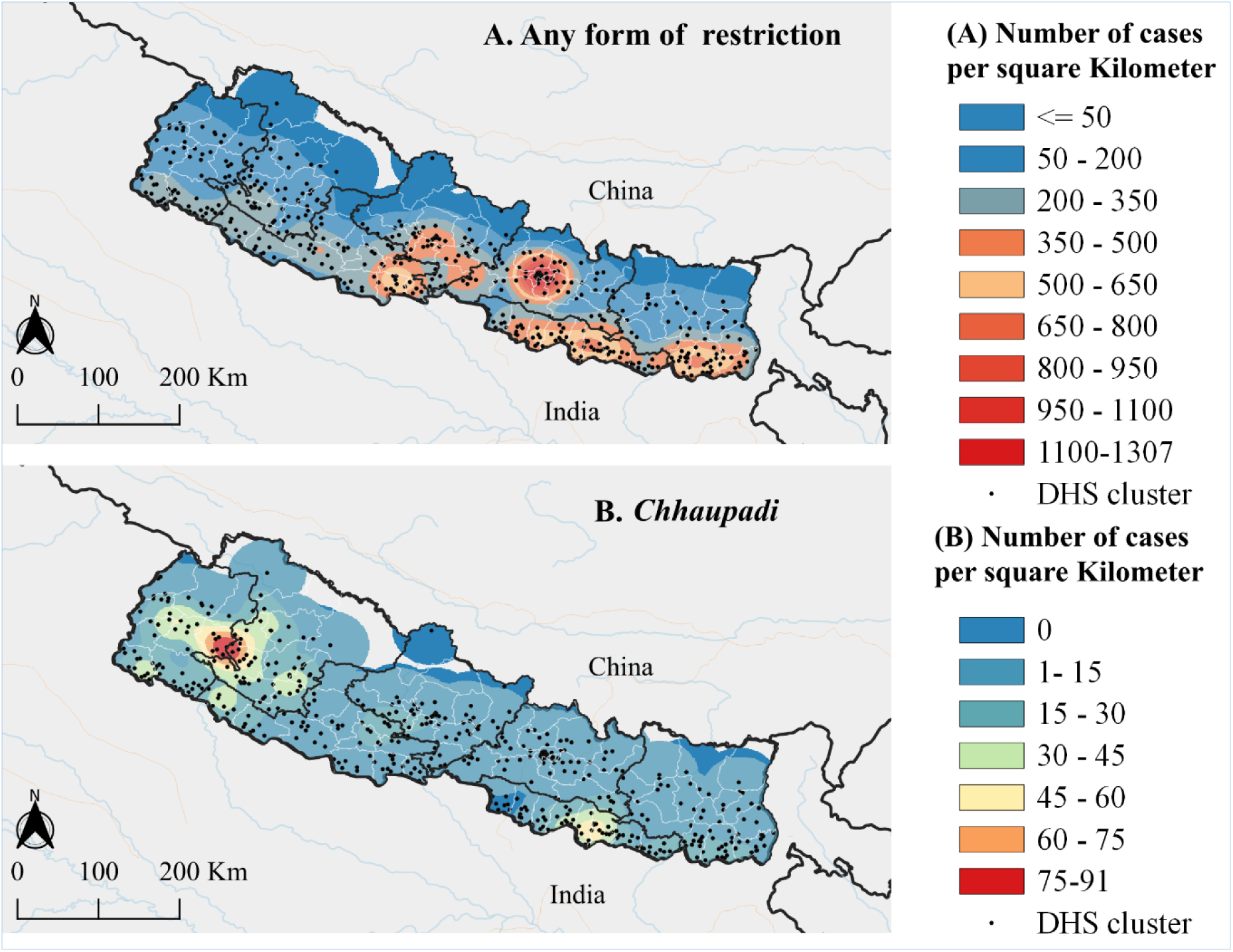
Continuous surface map showing the spatial case counts of individuals who experienced any form of menstrual restriction (panel A) and *Chhaupadi* (panel B), and the locations of demographic and health survey clusters.

Local cluster analyses identified specific areas in Sudurpashchim and Karnali as high-prevalence areas for all forms of menstrual restrictions. The prevalence clusters of *Chhaupadi* (**Figure 2**A) aligned with clusters of absolute case counts (**Figure 3**B), suggesting that targeting these high-prevalence areas will likely lead to a significant overall decline in *Chhaupadi* case counts across the country. However, this does not hold for the other types of menstrual restrictions, in which the case counts follow the population density distribution, with notable clusters observed in the urban setting of Kathmandu where the prevalence is relatively low (**Figure 2**A compared to **Figure 3**A).

A key finding of our study is the continued practice of *Chhaupadi* despite legislative interventions thus highlighting the intricate interplay between cultural norms and legal frameworks (Baumann, Lhaki, et al., 2021). This underscores the need to go beyond legislative measures and emphasis on culturally sensitive community engagement and active involvement of local and religious leaders in dismantling the deeply rooted harmful cultural norms (Baumann, Merante, et al., 2021; Sommer et al., 2016). The risk maps generated through the Gaussian kernel density estimation provide a visual representation of the geographical distribution of the various menstrual discriminatory practices. Armed with this spatial intelligence, policymakers and health practitioners can strategically allocate resources by tailoring interventions to the needs of each region. By recognizing the high-prevalence clusters as focal points allows for the development of interventions characterized by cultural sensitivity. These interventions may include targeted education programs and sensitization to challenge and transform the deeply rooted beliefs perpetuating menstrual stigma (Thapa et al., 2019).

Untargeted interventions, while well-intentioned, may fall short in areas where menstrual discriminatory practices are deeply ingrained. We contend that high-prevalence clusters, if left unaddressed, can serve to perpetuate the harmful cultural norm thus hindering the broader efforts to eliminate the vice across the country. We hypothesis that directing community-based interventions towards these specific clusters could be a highly effective way to break the cycle of harmful cultural believes and expedite progress towards a society where women can manage their menstrual health without facing discrimination. In doing so, we advocate for an approach that not only aims to eliminate discriminatory practices but also seeks to empower women, challenge harmful cultural believes, and promote an environment of inclusivity.

Women residing in households led by older household heads were associated with a higher likelihood of experiencing *Chhaupadi* or household level restrictions. This observation suggests a potential manifestation of conservative attitudes towards culture within the elderly demographic. In addition, women residing in households led by female household heads were associated with lower likelihoods of experiencing *Chhaupadi* or household level restrictions, suggesting a potential role of female leadership in challenging cultural norms. In keeping with previous findings, ethnicity/caste, and wealth of the household emerged as key determinants of menstrual restrictions (Baumann et al., 2019; Baumann, Lhaki, et al., 2021). Whilst richer households were associated with an increased likelihood of experiencing restrictions on religious activities, wealth was notably linked to protection against *Chhaupadi*. Furthermore, older women were associated with an increased likelihood of experiencing restrictions on religious activities, whilst education was associated with a heightened risk of menstrual restrictions within the household, but was not associated with *Chhaupadi*. Ethnicity/caste was associated with variations in all forms of menstrual restrictions, indicating the intersectionality of social demographic and cultural factors (Thapa et al., 2019).

Despite the robust statistical approaches used in this study, a substantial portion of the variability in the regression models remain unexplained. Therefore, there is need for additional research to identify the unmeasured determinants of menstrual discrimination that may act at various geographic scales. Understanding these unmeasured predictor variables is crucial for the development of comprehensive interventions that address the root cause of menstrual discrimination. Intervention designed against menstrual discriminatory practices should not only target high-prevalence clusters, as highlighted in our study, but also incorporate a research component to test their utility and cost effectiveness in real-world settings.

The significance of our study lies not only in its national scope but also in its novel approach to understanding the geographical nuances of menstrual health challenges in Nepal. Our study is the first to explore the micro-geographical variation of menstrual discriminatory practices against women aged 15–49 years. By delving into the geographical nuances of menstrual restrictions related to religious activities, household level and *Chhaupadi*, we aim to guide targeted interventions and resource allocation. Importantly, the estimation of case counts at 1 km^2^ spatial scale ensures that precise targeting can be done at any desired geographical scale.

Although our analysis provides a detailed description of menstrual discriminatory practices against girls and women in Nepal, we recognize some limitations. First, we could not establish whether during *Chhaupadi*, menstruating girls and women slept in make a shift-hut, cowshed, separate house within the homestead or separate room within the main house. Future studies may consider the various types of sleeping places during *Chhaupadi* in order to understand the evolving dynamics of the practice. Another limitation of our study is that we did not distinguish between internalized and externalized stigma. Lastly our analyses relied on reported data that may be subject to social desirability bias, however, regional estimates of prevalence are consistent with previous studies suggesting that the bias is likely minimal.

In conclusion, our study contributes valuable insights into the complexity of menstrual health challenges in Nepal, shedding light on the spatial patterns and determinants of menstrual discriminatory practices. By elucidating the geographical variations, our findings provide a foundation for targeted community based interventions such as the cultural behaviour change education (Amery et al., 2023; Thapa et al., 2019; Thomson et al., 2019). The identification and prioritization of vulnerable populations are crucial steps toward dismantling the deeply rooted beliefs and promoting a more inclusive and equitable society. As Nepal progresses towards sustainable development, addressing menstrual health disparities is imperative for achieving gender equality and societal well-being. Our study calls for sustained efforts, tailored community-based interventions, community engagement and policy initiatives to break the shackles of menstrual stigma and discrimination.

## Data Availability

All health data used in this analysis are available to download free of charge by registered users from the Demographic and Health Surveys Program. Registration, instructions on data access are available online at https://dhsprogram.com/data/new-user-registration.cfm. For data see https://dhsprogram.com/data/dataset_admin/download-manager.cfm. For full dataset access instructions see https://dhsprogram.com/data/Using-DataSets-for-Analysis.cfm.

https://dhsprogram.com/data/dataset_admin/download-manager.cfm

https://dhsprogram.com/data/Using-DataSets-for-Analysis.cfm

## Authors Contributors

GB and PM conceived the study. GB curated data. GB, SA, DM and PM designed the methods and conducted literature review. GB and PM did the formal analysis. GB, SA, DM and PM wrote the original draft of the Article. GB, SA, DM and PM reviewed and edited the Article.

**All authors have read, and confirm that they meet, ICMJE criteria for authorship.**

## Supplementary materials

**Supp. Table 1:**
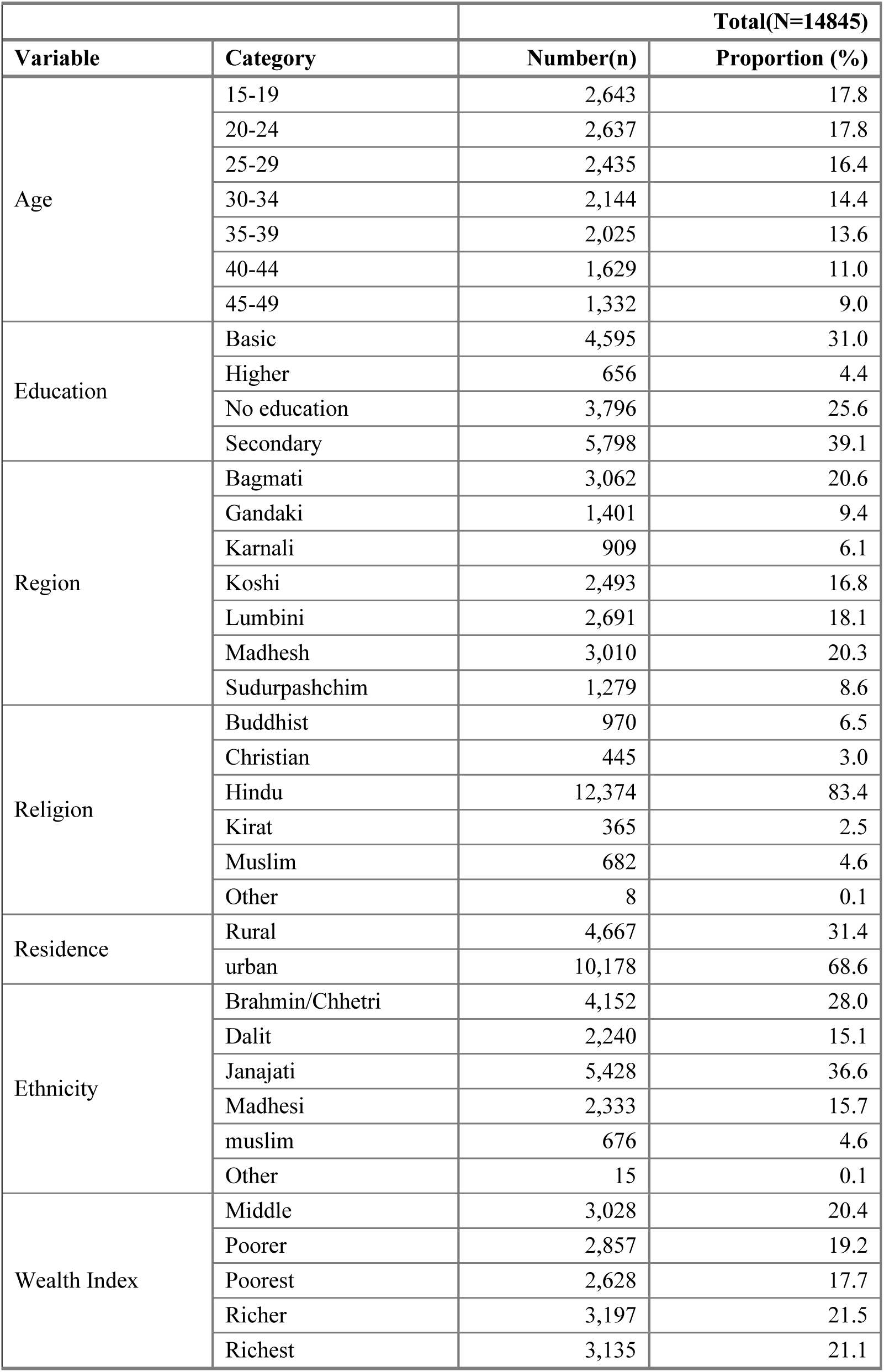
Social-demographic and economic characteristics of survey participants.

**Supp. Table 2:** Univariable Poisson regression of menstrual restrictions by social-demographic and economic factors.

|  | **Model1: Any restriction** |  |  | **Model2: Religious restrictions** |  |  | **Model3: Household level restrictions** |  |  | **Model4: Chhaupadi** |  |  |
| --- | --- | --- | --- | --- | --- | --- | --- | --- | --- | --- | --- | --- |
| **Characteristic** | **PR** | **95% CI** | **P-value** | **PR** | **95% CI** | **P-value** | **PR** | **95% CI** | **P-value** | **PR** | **95% CI** | **P-value** |
| **Age (ref: 15-19 years)** |  |  |  |  |  |  |  |  |  |  |  |  |
| 20-24 | 1.01 | 0.98-1.04 | 0.424 | 1.02 | 0.98-1.05 | 0.323 | 0.90 | 0.83-0.97 | 0.008 | 0.79 | 0.64-0.97 | 0.023 |
| 25-29 | 1.01 | 0.98-1.05 | 0.391 | 1.02 | 0.98-1.06 | 0.337 | 0.89 | 0.82-0.98 | 0.017 | 0.66 | 0.53-0.83 | <0.001 |
| 30-34 | 1.04 | 1.01-1.07 | 0.023 | 1.05 | 1.01-1.08 | 0.016 | 0.97 | 0.89-1.06 | 0.546 | 0.76 | 0.62-0.95 | 0.015 |
| 35-39 | 1.04 | 1-1.07 | 0.032 | 1.05 | 1.01-1.09 | 0.009 | 1.02 | 0.94-1.11 | 0.686 | 0.77 | 0.61-0.96 | 0.019 |
| 40-44 | 1.06 | 1.03-1.09 | <0.001 | 1.07 | 1.03-1.1 | <0.001 | 1.02 | 0.93-1.11 | 0.657 | 1.00 | 0.8-1.27 | 0.971 |
| 45-49 | 1.09 | 1.05-1.13 | <0.001 | 1.08 | 1.03-1.12 | 0.001 | 1.16 | 1.06-1.28 | 0.002 | 0.97 | 0.75-1.26 | 0.83 |
| **Highest level of education (ref: No education)** |  |  |  |  |  |  |  |  |  |  |  |  |
| Basic | 0.98 | 0.95-1.01 | 0.153 | 0.98 | 0.95-1.01 | 0.122 | 1.18 | 1.07-1.29 | 0.001 | 0.75 | 0.63-0.9 | 0.002 |
| Secondary | 1.01 | 0.98-1.04 | 0.61 | 1.01 | 0.98-1.04 | 0.484 | 1.35 | 1.22-1.5 | <0.001 | 0.69 | 0.58-0.84 | <0.001 |
| Higher | 1.08 | 1.04-1.12 | <0.001 | 1.11 | 1.06-1.16 | <0.001 | 1.64 | 1.43-1.9 | <0.001 | 0.37 | 0.21-0.66 | 0.001 |
| **Caste/Ethnicity (ref: Janajati)** |  |  |  |  |  |  |  |  |  |  |  |  |
| Brahmin/Chhetri | 1.26 | 1.22-1.31 | <0.001 | 1.19 | 1.15-1.24 | <0.001 | 3.82 | 3.4-4.28 | <0.001 | 8.47 | 5.87-12.22 | <0.001 |
| Dalit | 1.08 | 1.03-1.13 | 0.001 | 1.03 | 0.97-1.08 | 0.342 | 1.97 | 1.69-2.3 | <0.001 | 5.74 | 3.91-8.42 | <0.001 |
| Madhesi | 1.20 | 1.15-1.26 | <0.001 | 1.24 | 1.18-1.29 | <0.001 | 1.36 | 1.08-1.7 | 0.009 | 4.17 | 2.61-6.68 | <0.001 |
| Muslim | 0.96 | 0.84-1.11 | 0.598 | 0.97 | 0.83-1.13 | 0.718 | 0.42 | 0.25-0.7 | 0.001 | 0.00 | 0-0 | <0.001 |
| other | 1.13 | 0.88-1.46 | 0.33 | 1.18 | 0.92-1.52 | 0.198 | 2.93 | 1.53-5.6 | 0.001 | 7.60 | 1.73-33.39 | 0.007 |
| **Residence (ref: Rural)** |  |  |  |  |  |  |  |  |  |  |  |  |
| Urban | 1.03 | 0.99-1.07 | 0.127 | 1.06 | 1.02-1.11 | 0.009 | 0.95 | 0.83-1.08 | 0.423 | 0.53 | 0.39-0.71 | <0.001 |
| **Region/Province (ref: Bagmati)** |  |  |  |  |  |  |  |  |  |  |  |  |
| Gandaki | 1.02 | 0.94-1.1 | 0.682 | 0.98 | 0.89-1.08 | 0.678 | 1.09 | 0.88-1.34 | 0.421 | 1.94 | 1.07-3.51 | 0.029 |
| Karnali | 1.16 | 1.1-1.23 | <0.001 | 1.11 | 1.03-1.18 | 0.003 | 1.89 | 1.63-2.19 | <0.001 | 14.07 | 8.71-22.73 | <0.001 |
| Koshi | 1.09 | 1.03-1.16 | 0.004 | 1.11 | 1.03-1.18 | 0.003 | 1.14 | 0.95-1.37 | 0.173 | 1.77 | 0.93-3.38 | 0.082 |
| Lumbini | 1.06 | 1-1.13 | 0.05 | 1.05 | 0.99-1.13 | 0.121 | 1.10 | 0.91-1.32 | 0.333 | 2.17 | 1.18-3.97 | 0.012 |
| Madhesh | 1.09 | 1.02-1.16 | 0.008 | 1.13 | 1.06-1.21 | <0.001 | 0.44 | 0.34-0.57 | <0.001 | 2.55 | 1.48-4.37 | 0.001 |
| Sudurpashchim | 1.21 | 1.15-1.27 | <0.001 | 1.04 | 0.97-1.12 | 0.224 | 1.99 | 1.67-2.37 | <0.001 | 10.55 | 6.33-17.61 | <0.001 |
| **Gender of the head of the household (ref: Male)** |  |  |  |  |  |  |  |  |  |  |  |  |
| Female | 0.97 | 0.95-1 | 0.02 | 0.98 | 0.95-1 | 0.059 | 0.87 | 0.81-0.94 | <0.001 | 1.12 | 0.95-1.32 | 0.196 |
| **Age of the head of the household (ref: 15-24 years)** |  |  |  |  |  |  |  |  |  |  |  |  |
| 25-34 | 1.04 | 0.98-1.09 | 0.198 | 1.04 | 0.98-1.1 | 0.205 | 1.17 | 0.98-1.4 | 0.089 | 0.98 | 0.68-1.42 | 0.934 |
| 35-44 | 1.05 | 0.99-1.11 | 0.112 | 1.05 | 0.99-1.11 | 0.118 | 1.43 | 1.2-1.71 | <0.001 | 1.18 | 0.81-1.73 | 0.388 |
| 45-54 | 1.07 | 1.01-1.13 | 0.017 | 1.07 | 1.01-1.14 | 0.028 | 1.51 | 1.26-1.81 | <0.001 | 1.10 | 0.75-1.62 | 0.617 |
| 55-64 | 1.09 | 1.03-1.16 | 0.002 | 1.08 | 1.02-1.15 | 0.013 | 1.66 | 1.39-1.99 | <0.001 | 1.08 | 0.74-1.58 | 0.693 |

|  | Model1: Any restriction |  |  | Model2: Religious restrictions |  |  | Model3: Household level restrictions |  |  | Model4: Chhaupadi |  |  |
| --- | --- | --- | --- | --- | --- | --- | --- | --- | --- | --- | --- | --- |
| Characteristic | PR | 95% CI | P-value | PR | 95% CI | P-value | PR | 95% CI | P-value | PR | 95% CI | P-value |
| >=65 | 1.10 | 1.04-1.17 | 0.001 | 1.07 | 1-1.14 | 0.055 | 1.89 | 1.57-2.26 | <0.001 | 1.34 | 0.91-1.98 | 0.141 |
| **Wealth Index (ref: Poorest)** |  |  |  |  |  |  |  |  |  |  |  |  |
| Middle | 1.02 | 0.98-1.07 | 0.333 | 1.14 | 1.08-1.2 | <0.001 | 0.66 | 0.58-0.76 | <0.001 | 0.40 | 0.29-0.55 | <0.001 |
| Poorer | 1.04 | 0.99-1.08 | 0.104 | 1.13 | 1.07-1.19 | <0.001 | 0.71 | 0.63-0.8 | <0.001 | 0.44 | 0.33-0.58 | <0.001 |
| Richer | 1.04 | 0.99-1.09 | 0.107 | 1.15 | 1.09-1.22 | <0.001 | 0.69 | 0.6-0.8 | <0.001 | 0.28 | 0.2-0.4 | <0.001 |
| Richest | 1.06 | 1.01-1.12 | 0.018 | 1.19 | 1.13-1.26 | <0.001 | 0.87 | 0.76-1 | 0.044 | 0.13 | 0.08-0.2 | <0.001 |

